# COVID-19 Severity Index: A predictive score for hospitalized patients

**DOI:** 10.1101/2020.08.12.20166579

**Authors:** Iván Huespe, Indalecio Carboni Bisso, Nicolas Alejandro Gemelli, Sergio Terrasa, Sabrina Di Stefano, Valeria Burgos, Jorge Sinner, Marcelo Risk, Eduardo San Román, Marcos Las Heras

## Abstract

**Introduction:** Pandemics pose a major challenge for public health preparedness, requiring a coordinated international response and the development of solid containment plans. An early and accurate identification of high-risk patients in the course of the actual COVID-19 pandemic is vital for planning and for making proper use of available resources.

**Objective:** The purpose of this study was to identify the key variables to create a predictive model that could be used effectively for triage.

**Methods:** A narrative literature review of 651 articles was conducted to assess clinical, laboratory and imaging findings of COVID-19 confirmed cases. After screening, 10 articles met the inclusion criteria and a list of suggested variables was gathered. A modified Delphi process analysis was performed to consult experts in order to generate a final list of variables for the creation of the predictive model.

**Results:** The modified Delphi process analysis identified 44 predictive variables that were used for building a severity prediction score, the *COVID-19 Severity Index*.

**Conclusion:** Specifically designed for current COVID-19 pandemic, *COVID-19 Severity Index* could be used as a reliable tool for strategic planning, organization and administration of resources by easily identifying hospitalized patients with higher risk of transfer to Intensive Care Unit (ICU).

## INTRODUCTION

Infectious disease outbreaks constitute a serious problem to global health with a major impact on countries’ economies, its healthcare systems and resources. The spread of Severe acute respiratory syndrome coronavirus 2 (SARS-CoV-2) known as COVID-19, has already gone into pandemic proportions registering, at the moment of this study, a total of 20 million confirmed cases, 736.000 deaths and 13 million recovered patients across 215 countries in a short elapse of time. The way in which outbreaks affect countries depends on multiple factors and its impact is difficult to foresee. However, the numbers of infected people and casualties are evidence that despite the attempts to plan in advance, the global healthcare systems remain unprepared^1^.

The intensity of staffing needed and the sophisticated training required for the care of patients with viral infections during pandemics result in the fact that a relatively small number of patients could easily overwhelm healthcare systems.

An accurate identification of variables related to worse outcome is key for triaging and adapting intensity of the care that each patient requires. The previous would allow an effective strategic planning and a better administration of human and material resources. Moreover, the need for a sensitive and predictive model is mandatory to avoid a delayed recognition of severely ill patients or even those at risk of presenting further complications. During the early phase of COVID-19 pandemic *Liao et al*. propose an early warning score based on an adapted version of the National Early Warning Score 2 (NEWS-2) adding age as a variable to reflect emerging evidence of age as an independent risk factor for survival ^2^. In that score, patients were divided into four categories based on the risk of respiratory failure: low, medium, high, and extreme. The score was used to manage the monitoring frequency and to activate a rapid response team.

Early Warning Scores (EWS) predict deterioration in hospitalized patients ^3^, but are designed for general hospitalized patients in a non-pandemic scenario. COVID-19 pandemic has now a high proportion of hospitalized patients with a single disease. Therefore, a specific EWS for COVID-19 including laboratory test results, clinical features and radiological findings, could improve the detection of high-risk patients in the aim of optimizing the management of hospital resources. The previous is key in low-income countries where resources are insufficient even before affronting this pandemic.

Aware of the impact of the current pandemic, *“COVID-19 Severity Index*” is developed as a triage tool based on the NEWS-2 score, that could rapidly and reliably be used by frontline healthcare personnel to identify high-risk patients.

## METHODS

### Protocol registration

This project was approved by the Ethics Committee for Research Protocols at Hospital Italiano de Buenos Aires (Cod. 1290). Voluntary completion of the questionnaire implied consent and the participants’ responses were received and analyzed anonymously.

### Methodological approach

A narrative review was conducted to generate a list of possible predictors based on clinical signs and symptoms, comorbidities, laboratory and radiographic findings. After initial identification of predictive variables, they were subjected to expert analysis through a 2-round Delphi process ^4^. The output was a set of potential variables based on expert opinion to be added to the NEWS-2.

### Narrative review

Narrative review was conducted in April 2020 using the Ovid MEDLINE and medRxiv database for articles written in English and published until April 2020. The search strategy included terms such as “COVID-19”, “Risk Factors”, “Respiratory Insufficiency” and “Mortality”. Studies were selected on the base of the following inclusion criteria: population over 18 years of age where signs and symptoms were recorded together with comorbidities, laboratory and radiographic findings and in which all these parameters were valued against the occurrence of death or disease severity in confirmed COVID-19 patients.

### Delphi process

Delphi method is a process used to arrive at a true consensus by surveying a panel of experts who answer several rounds of questionnaires. Responses are aggregated and shared with the group after each round, allowing experts to adjust their answer based on their interpretation of the “group response” provided. Ultimate result is meant to be a true consensus of what the group thinks.

A 2-round Delphi process was started in April 2020 and completed in May 2020. It enabled the construction of a list of 44 predictive variables identified through the narrative review that could be used as criteria for the selection of patients with worse clinical outcome. This was achieved using feedback from 14 expert contributors with diverse backgrounds and located in different countries affected by COVID-19.

### Participants

Experts from both resources-rich and resource-limited settings were recruited, including professionals from Argentina, Chile and Canada. Participants are involved in the care of critically ill adults and active in medical research areas including critical care, infectious diseases and pneumology on a daily basis. First contact was by email to communicate the objective of the study and extend invitation to participate in both rounds of the Delphi process. The target sample size of expert contributors was 14.

### Round 1

The first round of the Delphi process had a 7 day elapse; it was initiated on April 15th of 2020 and completed by April 22nd of 2020. The questionnaire form was distributed among expert participants who were asked to evaluate the variables gathered from the literature review before mentioned and suggested as potential predictors of worse outcome in COVID-19 patients.

When the participant answered “yes” to the fact that a given variable was valuable as a predictor, they were prompted to evaluate 3 domains: predictive potential, measurement reliability, level of training and/or resources required to measure and collect the variable ^4^ Each domain had 4 possible answers: high (3), moderate (2), minimal (1), or not applicable (0). Finally there was an option for the participant to make comments regarding each variable. Moreover, they were encouraged to add new variables to the suggested set.

At the stage of analysis, each answer was given a number between 3 and 0 (high (3), moderate (2), minimal (1), not applicable (0)) based on the strength of the response. The numbers for each domain were tabulated to calculate a weighted effect (WE) to help determine the selection threshold. The WE was calculated following the formula exemplified below, that doubles the weight of the value for predictive potential, adds the value for measurement reliability and subtracts the value for level of resources and/or training required.

> **Weighted Effect** = Predictive Potential x2 + Reliability - Resources or Training
>
> eg.: **Weighted Effect**_Asthma_ = Moderate (2) x2 + Moderate (2) - Minimal (1)
>
> **Weighted Effect**_Asthma_ = 5

WE was calculated for each variable valued by each expert opinion. The sum of the WE’s for a given variable was ranked for further selection of those with the greatest value achieved whereas WE of variables considered as medical records, were analyzed separately from clinical features.

Afterwards, a threshold was chosen based on a desired number of predictors. Then variables which WE scored above the threshold, were included in a final set of predictor variables. Those variables that were below the threshold, were carefully reviewed by the research team and discharged or included in Round 2 for re-evaluation depending on the value obtained. Any additional variables proposed by participants were evaluated in Round 2 when they were considered clinically distinct from the variables already assessed in Round 1.

### Round 2

The 2nd Round was conducted in a 7 day elapse of time, from 29 April of 2020 to 7 May of 2020. It consisted in re-evaluating selected variables from Round 1 as well as evaluating newly suggested variables. Participants were provided the responses from Round 1, and a threshold procedure similar to the one used in the initial round was used in the second round. Each round demanded 7 days for completion given to each expert consulted who was sent a reminder one day before the deadline. The final set of variables was obtained and results were analyzed as follows.

### Development of the questionnaire with Google Forms

Questionnaire was developed using the secure web-based application Google Forms. Participant’s own responses from Round 1 were available in Round 2 along with a mean response from the other participants.

## RESULTS

### Narrative review

After analysis, ten articles fulfilled the final inclusion criteria and were, therefore, considered. Sixty-four relevant variables predictors analyzed in these studies were summarized to generate a master list (***Supplementary material* 1**) created using Microsoft Excel to keep track of each predictor variable and the frequency of repetition in other studies as a presumed indicator of its predictive potential and relative commonality. Each variable was organized into the following categories: 1) patient’s characteristics; 2) signs and symptoms; 3) scores; 4) laboratory findings; 5) chest x-ray findings; 6) comorbidities. All variables were presented at either binary or continuous variable, depending on the way in which it was presented in the original study.

### Round 1

There was a high level of agreement on the following variables alerting to a worse outcome: age, male gender, dyspnea, d-dimer > 1 μg/ml, lymphopenia, “Sequential Organ Failure Assessment” (SOFA) score, bilateral compromise in chest x-ray and comorbidities such as chronic heart failure, diabetes with end-organ damage and hypertension. The thresholds for age and lymphopenia were discussed in Round 2.

Additionally, there was a high level of agreement to not include: pregnancy, plasma albumin, pro-B-type natriuretic peptide, lactate dehydrogenase and other comorbidities such as chronic obstructive pulmonary disease (COPD), asthma, chronic renal disease, solid tumor, tuberculosis, active smoking and diabetes without end-organ damage.

Since there was a moderate level of agreement regarding thrombocytopenia, C reactive protein, creatinine serum levels, chest x-ray findings other than bilateral compromise, those variables were discussed in Round 2.

Experts also proposed 2 new variables; findings in pulmonary ultrasound and reticulonodular interstitial pattern in the chest x-ray. These 2 new variables were not considered due to interobserver variability and the need for highly trained professionals to perform and to interpret those studies.

### Round 2

Due to the lack of a specific cut-off value for age and lymphopenia, possible thresholds were proposed to participants. After Round 2, cut-off values for age were defined as follows: low risk for age < 60; moderate risk for ages between 60 and 65; and high risk for age > 65. Regarding lymphopenia, thresholds were: >1000 mm^3^, low risk; between 500 and 1000 mm^3^, moderate risk; and less than 500 mm^3^, high risk.

Variables from Round 1 that did not reach the WE threshold value for immediate consideration, were reassessed. Low platelet count, C-reactive protein, lactate dehydrogenase, and serum creatinine were reconsidered to select the one with a higher predictive value of the worse outcome. Consensus on platelet count <100000 mm^3^ was the variable with higher potential. List of all potential variables proposed to the Delphi process are available for reference in ***Supplementary material 2***. The final set variables selected from the Delphi process are exposed in **Figure 1**.

**Figure 1.**
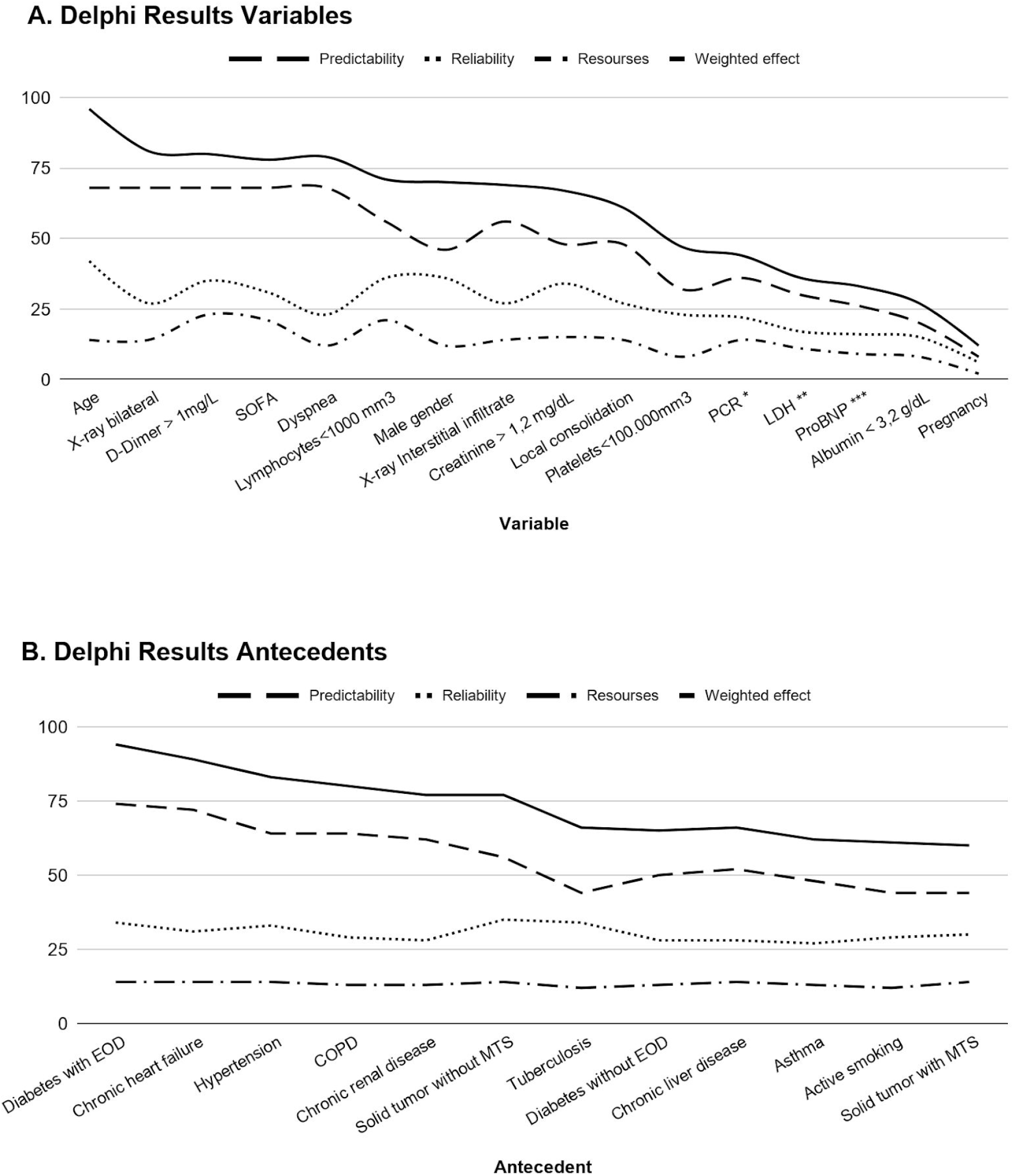
Delphi process results. In the first round of the Delphi process, variables were suggested to participants to determine their opinion as to its value as a predictor of worse outcome in COVID-19 patients. If the participant responded “yes”, they were prompted to evaluate the variable according to three domains: predictive potential, measurement reliability, and the level of training and/or resources required to measure and collect the variable. Each domain consisted of 4 possible answers: high (3), moderate (2), minimal (1), or not applicable (0). For the analysis, each answer was given a number between 0 and 3 based on the strength of the response. The sum for each domain was tabulated to calculate a weighted effect (WE) to help determine the selection threshold. The WE was calculated by doubling the weight of the value for predictive potential, adding the value of measurement reliability and subtracting the value of level of resources and/or training required. The WE was calculated for each variable by expert opinion. The sum of the WE for each variable was ranked for further selection of those with the greatest value. A threshold was chosen based on the desired number of predictors. Variables that scored a WE above the threshold were included in the final set of predictor variables. Predictors below the threshold were carefully reviewed by the research team and were discharged or included in Round 2 for re-evaluation, depending on the value obtained. **A**. The trends of the sum of responses to each domain for all 16 variables evaluated in Round 1. **B**. The trends of the sum of responses to each domain for all 12 antecedents evaluated in Round 1. COPD: chronic obstructive pulmonary disease; EOD: end-organ damage; MTS: metastasis.PCR: Protein C reactive; LDH: Lactate dehydrogenase *Values of PCR, proposed were: >10, >100 or >200 mg/dl **Values of LDH, proposed were: > 250, >300 or >350 U/L ***Values of Pro B-type Natriuretic Peptide, proposed were > 350, >500 or >1000 pg/mL

### Final Score

Following the analysis and the application of the modified Delphi process, a final set of selected variables was combined with a modified NEWS-2 score to generate the *COVID-19 Severity Index*. The modifications to NEWS-2 Score are described as follows. In the case that the patient needs supplemental oxygen, he will receive 3 score-points instead of 2. Addition of 1 and 2 score-points related to low blood pressure was eliminated. Low temperature only added 1 score-point if it was less than 35.6°C instead of 36,1°C, while 1 point-score was added if the temperature was 38°C or higher. *COVID-19 Severity Index* score is exposed in ***Table 1***. Patients were divided into four risk categories based on their score (***Table 2***).

**Table 1.**
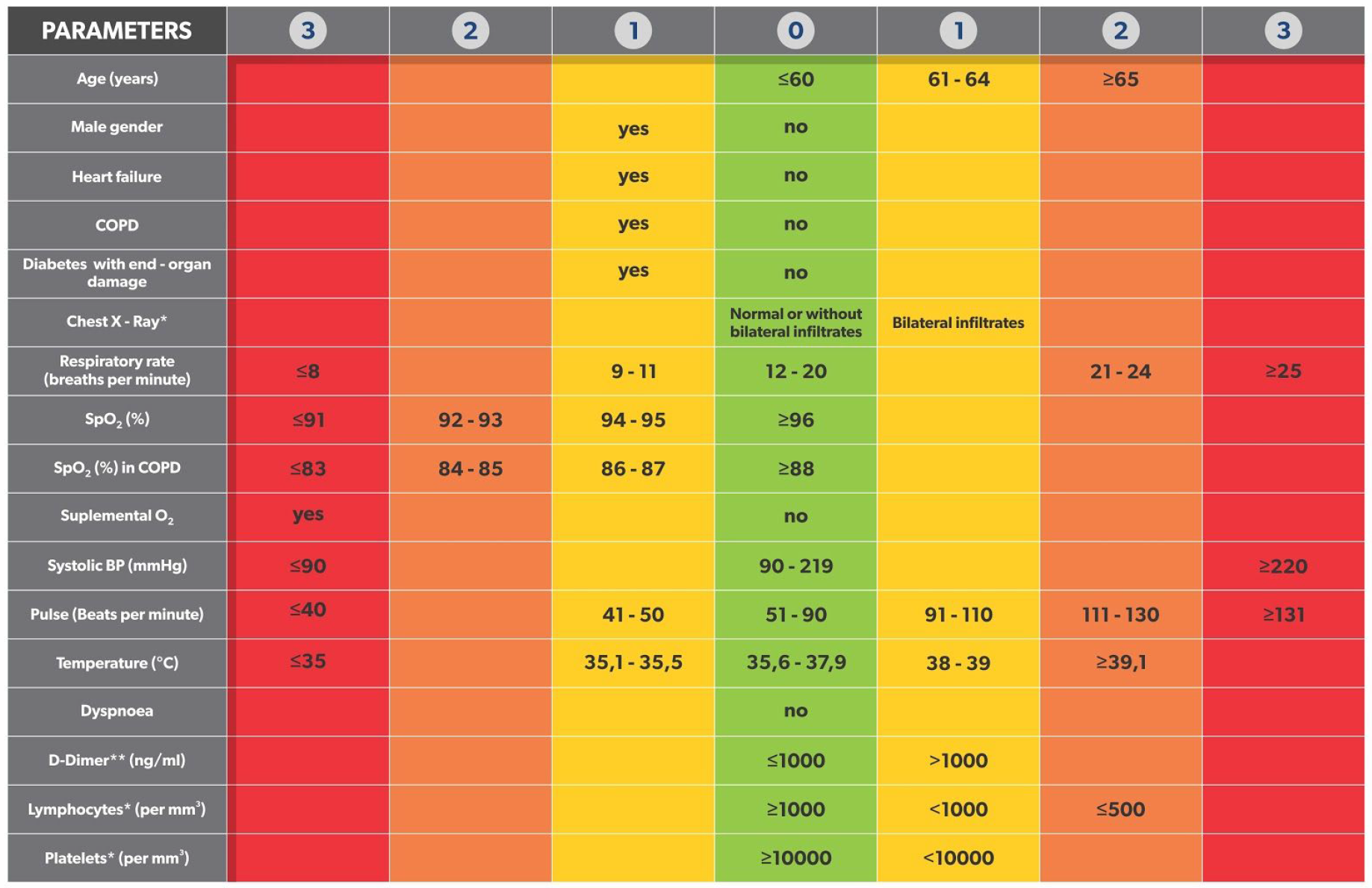
COVID-19 Severity Index. * Chest X-Ray should be analyzed on admission but it will be reconsidered when a new one is performed. ** If laboratory test results have more than 48 hours, they will not be considered. COPD: chronic obstructive pulmonary disease.

**Table 2.**
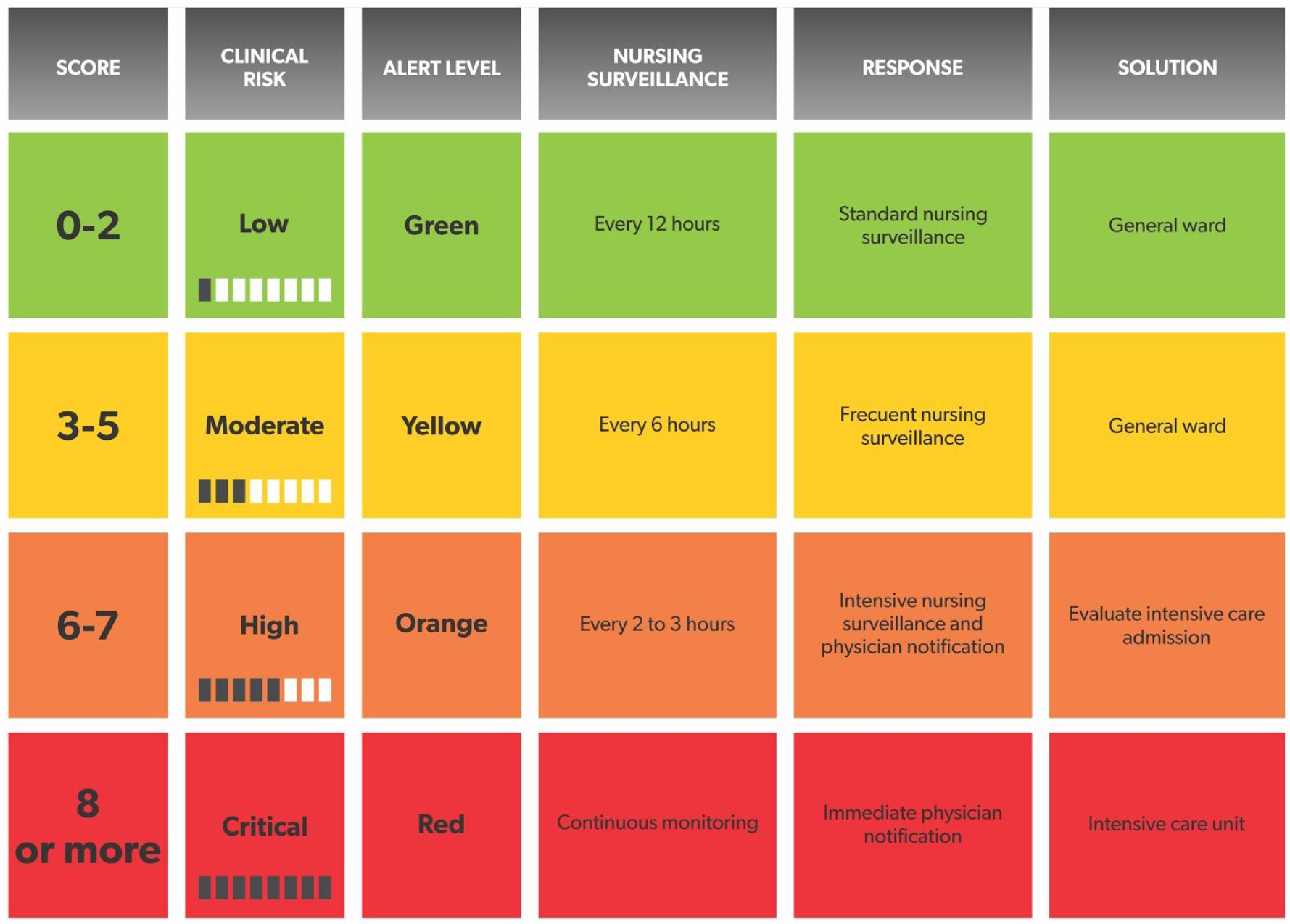
COVID-19 Severity Index risk chart.

| SCORE | CLINICAL RISK | ALERT LEVEL | NURSING SURVEILLANCE | RESPONSE | SOLUTION |
| --- | --- | --- | --- | --- | --- |
| 0-2 | Low | Green | Every 12 hours | Standard nursing surveillance | General ward |
| 3-5 | Moderate | Yellow | Every 6 hours | Frequent nursing surveillance | General ward |
| 6-7 | High | Orange | Every 2 to 3 hours | Intensive nursing surveillance and physician notification | Evaluate intensive care admission |
| 8 or more | Critical | Red | Continuous monitoring | Immediate physician notification | Intensive care unit |

### Validation

Prediction capacity of this score was studied to evaluate its predictive potential of ICU transfer in 24 and 48 hours elapse of time.

A group of 220 patients with confirmed COVID-19 were evaluated; 19 of which were unexpectedly transferred to ICU; and 17 of which were transferred to ICU during the first 3 days, one at 5th day and another at 6th day of hospitalization.

A comparison between *COVID-19 Severity Index*, NEWS score adapted by Liao et al. ^2^ and NEWS-2 score was made. All three scores were measured on the first, second and third day after hospital admission of the patients.

For those patients who were initially admitted into general wards and were later transferred to the ICU, the score was retrospectively applied for the 72, 48 and 24 hours prior to the ICU admission, with the intention to identify whether they were parameters that could predict the need of a more intensive monitoring.

A comparative analysis of the area under the curve (AUC) for the different scores evidenced a better capacity of the *COVID-19 Severity Index* to predict the need for ICU admission. When applied in the 24 hours prior to ICU admission, the AU-ROC for the score *COVID-19 Severity Index* was 0.94 *vs*. 0.88 for the modified NEWS score developed by *Liao et al*., and 0. 80 for NEWS-2 (***Figure 2***). When applied in the 48 hours prior to ICU admission, the AU-ROC for *COVID-19 Severity Index* was 0.88, for the modified NEWS was 0.84 and 0.62 for NEWS-2.

**Figure 2.**
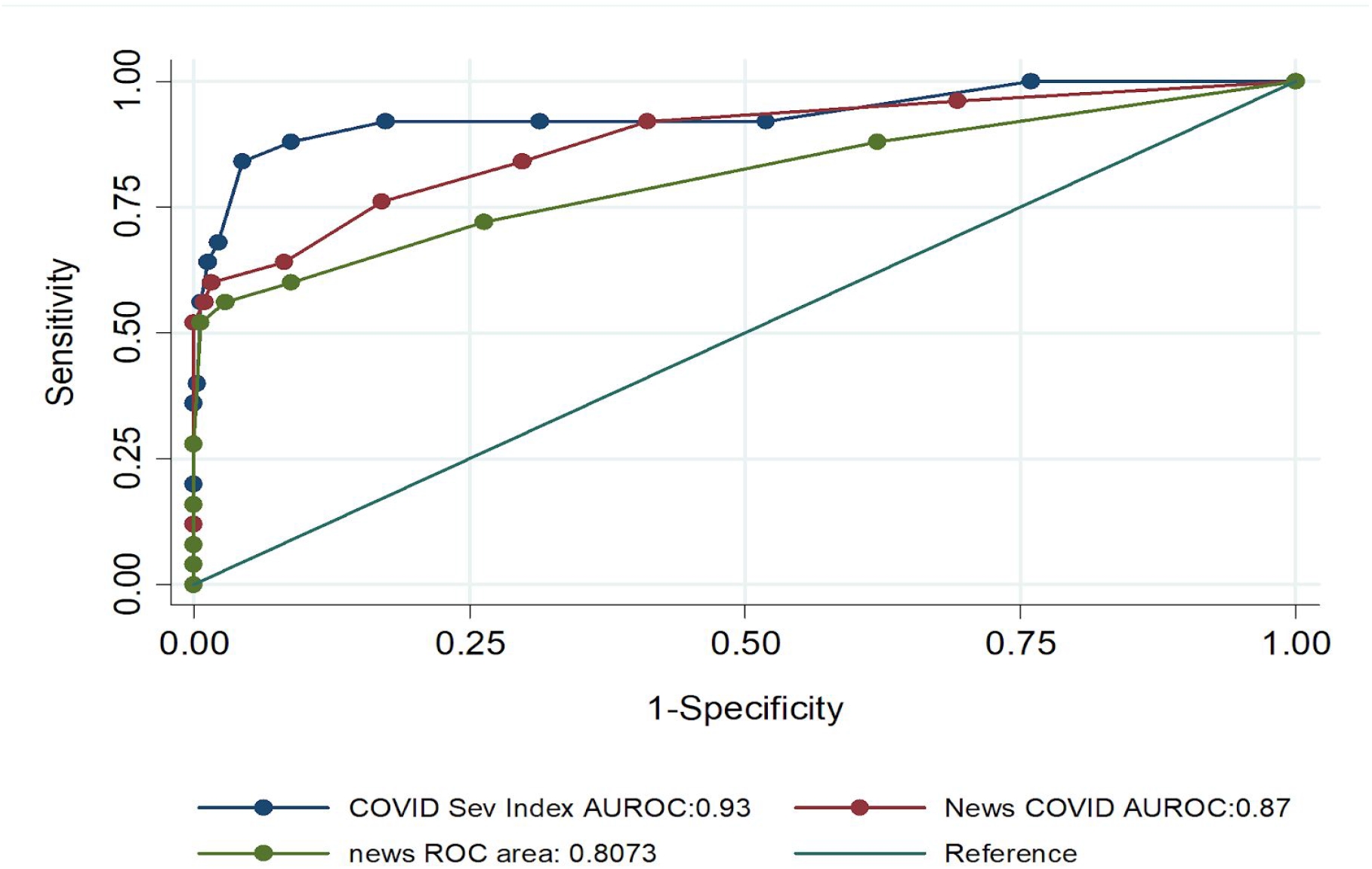
Area Under the Curve of COVID Severity Index, NEWS COVID proposed by Liao et al.^2^ and NEWS-2 Score to predict unexpected ICU transfer in the next 24 hours.

## DISCUSSION

In this study, an EWS was designed to predict progression towards critical illness among COVID-19 infected patients during hospitalization.

Although NEWS-2 score is the mainly used EWS, there are few published studies on it’s use in the specific context of COVID-19 ^5^. A paper published during the early phase of the COVID-19 pandemic offered an EWS based on an adapted version of the NEWS-2 score in which age > 65 years was added to reflect emerging evidence of age as an independent risk factor for survival ^2^.

The complexity of COVID-19 and the multiple variables involved in its course evidenced the need to search for a more specific score that could be used in this single disease to better discern among patients at risk of presenting severe infection. The development and use of a simple tool built on the basis of signs and symptoms with moderate to strong predictive potential, could ideally facilitate the triage process and expedite the care for hospitalized adult patients with COVID-19.

Due to the current lack of evidence, the effort to carry out careful research with a methodologically solid process was paramount. The narrative review allowed the research team to balance information from peer-reviewed articles as well as urgent data reported in preprints.

Additionally the Delphi method allowed to merge clinical expertise to theoretical reasoning. Delphi process was chosen with the objective of achieving consensus among a panel of experts on a defined issue, using iteration of a questionnaire and aggregating the answers to provide feedback to the participants after each completed round. This method as a way of generating consensus is widely applied in diverse fields such as program planning and resource assessment, even in the healthcare sector ^6^. One of the advantages of using this method is the facilitation of consensus-building online which significantly enabled the participation of subject matter experts from various locations worldwide in the current pandemic scenario ^4^

The 10 variables added to the modified NEWS-2 score required for calculation of the risk of developing a critical illness, are usually available at Hospital admission. In addition to the previous, a web-based calculator was created to facilitate the use of the tool. *COVID-19 Severity Index* is a dynamic tool designed to be actualized with the clinical changes of the patient, in the aim of detecting clinical deterioration 24 to 48 hours prior to ICU transfer.

Even though *COVID-19 Severity Index* has a short scale validation, it was designed by experts’ opinions. It is the case that opinions may be impacted by experts’ training, exposure, and expertise but this added biological plausibility. The application of this score is being carried out in patients at Hospital Italiano de Buenos Aires to define the intensity of nursing monitoring required.

## CONCLUSION

Specifically designed for the current COVID-19 pandemic, *COVID-19 Severity Index* could serve as a reliable tool for strategic planning, organization and administration of resources by easily distinguishing hospitalized patients with higher risk of ICU transfer.

## Data Availability

All Data is Available.

## Acknowledgments

We would like to thank the experts Marcos Marino, Sergio Giannasi, Fernando Vazquez, Martin Hunter, Leonardo Uranga, Chung Kyu, José Dianti, Manuel Tisminetzky, Bruno Ferreyro, Federico Angriman, Eduardo De Vito, Silvia Quadrelli, Alejandro Chirino, Eduardo Tobar and Pablo Gastaldi for their enthusiastic contribution and suggestions in the modified Delphi process.

Also, we would like to thank Lisandro Ziperovich (*Zipper Art*) for the artwork of this study and Maria de los Angeles Magaz for draft editing.

## Data Availability Statement

all relevant data are within the manuscript and its Supporting Information files.

## Author Contributions

Dr. Huespe had full access to all data in the study and takes responsibility for integrity of the data and accuracy of the data analysis.

<u>Conceptualization:</u> Huespe, Carboni Bisso

<u>Formal analysis</u>: Huespe, Carboni Bisso, Terrasa

<u>Methodology</u>: Huespe, Carboni Bisso, Terrasa

<u>Project administration</u>: Huespe, Carboni Bisso, Las Heras

<u>Visualization</u>: Las Heras, Risk, Terrasa

<u>Writing – original draft</u>: Gemelli

<u>Writing – review & editing</u>: Huespe, Carboni Bisso, Gemelli

<u>Administrative, technical, or material support:</u> San Román, Las Heras, Di Stefano, Burgos

<u>Supervision:</u> San Román, Las Heras, Risk, Sinner

